# MODELLING OF COVID-19 OUTBREAK INDICATORS IN CHINA BETWEEN JANUARY AND APRIL

**DOI:** 10.1101/2020.04.26.20080465

**Authors:** Senol Çelik, Handan Ankarali, Ozge Pasin

## Abstract

**Background:** The aim of this study is to explain the changes of outbreak indicators for coronavirus in China with nonlinear models and time series analysis. There are lots of methods for modelling. But we want to determine the best mathematical model and the best time series method among other models.

**Methods:** The data was obtained between January 22 and April 21, 2020 from China records. The number of total cases and the number of total deaths were used for the calculations. For modelling Weibull, Negative Exponential, Von Bertalanffy, Janoscheck, Lundqvist-Korf and Sloboda models were used and AR, MA, ARMA, Holt, Brown and Damped models were used for time series. The determination coefficient (R^2^), Pseudo R^2^ and mean square error were used for nonlinear modelling as criteria for determining the model that best describes the number of cases, the number of total deaths and BIC (Bayesian Information Criteria) was used for time series.

**Results:** According to our results, the Sloboda model among the growth curves and ARIMA (0,2,1) model among the times series models were most suitable models for modelling of the number of total cases. In addition Lundqvist-Korf model among the growth curves and Holt linear trend exponential smoothing model among the times series models were most suitable model for modelling of the number of total deaths. Our time series models forecast that the number of total cases will 83311 on 5 May and the number of total deaths will be 5273.

**Conclusions:** Because results of the modelling has providing information on measures to be taken and giving prior information for subsequent similar situations, it is of great importance modeling outbreak indicators for each country separately.

## Introduction

Coronavirus outbreak (COVID-19) was first appeared in the Wuhan city, Hubei Province, in China. The virus was identified in the first half of January 2020 [1]. The epidemiological features of the disease are still unknown, and the number of total cases and the number of total deaths varies day by day. When the rapid spread and serious consequences of the disease were observed, precautions were taken and positive cases began to be recorded after the second half of January. Because the number of total cases and the number of total deaths are used for examining the course of the outbreak, modelling of these indicators are the important issue. The model results are valuable for determining the appropriate preventions. In the literature, it was observed that some models were used for short-term forecasts considering the previous data. It is possible to reduce the number of total cases and total deaths by taking necessary precautions. At this point, it is very important to make future estimates.

In this study, it was aimed to examine the course of outbreak in China with alternative statistical models by taking into the number of total cases and the number of total deaths for the day it became clear until April 20 and also we want to select the best model for defining with model selection criteria. For modelling Weibull, Negative Exponential, Von Bertalanffy, Janoscheck, Lundqvist-Korf and Sloboda models were used and AR, MA, ARMA, Holt, Brown and Damped models were used for time series.

## Materials and Methods

### Data

The data was obtained between January 22 and April 21, 2020 in China records. The number of total cases and the number of total deaths were used for the calculations. The reasons for choosing China for modeling are that, it is the first country to fight the outbreak, therefore it is possible to observe the natural course of the outbreak and it is the country that struggles for the longest time.

The number of total cases and the total deaths of coronavirus disease in the world were recorded daily by following the web address https://www.worldometers.info/coronavirus/.

### Models for describing the course of the outbreak

For coronavirus outbreak, the change in the number of total cases and the number of total deaths over time was analyzed separately. In the analysis of time-total case and time-total deaths, six different nonlinear models were used including Weibull, Negative exponential, Von Bertalanffy, Janoscheck, Lundqvist-Korf and Sloboda models (Table 1) [2]. For time series analysis, Box-Jenkins and exponential smoothing methods were used (Table 2 and Table 3).

**Table 1.** Growth Curves

| Model | Equation |
| --- | --- |
| Weibull model | $Y_t = A - be^{-kt^\gamma}, t \geq 0$ |
| Negative exponential model | $Y_t = A(1 - e^{-kt}), t \geq 0$ |
| Von Bertalanffy model | $Y_t = A(1 - be^{-kt})^3, t \geq 0$ |
| Janoscheck model | $Y_t = A(1 - be^{-kt^c}), c > 1$ |
| Lundqvist-Korf model | $Y_t = Ae^{-kt^{-d}}$ |
| Sloboda model | $Y_t = Ae^{-be^{-kt^\gamma}}, t \geq 0$ |

**Table 2:**
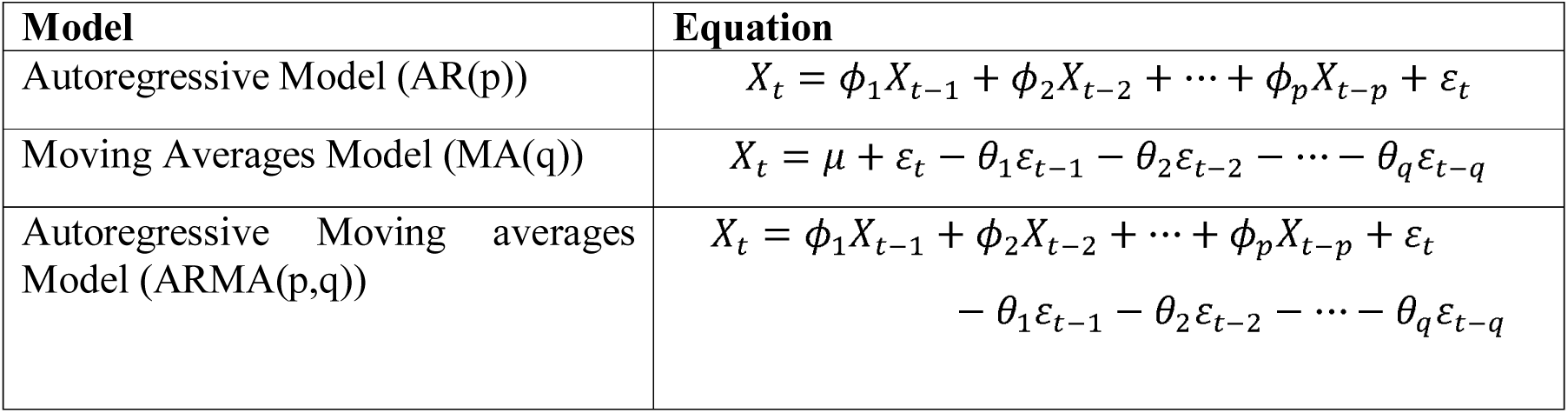
Box-Jenkins Models

**Table 3.**
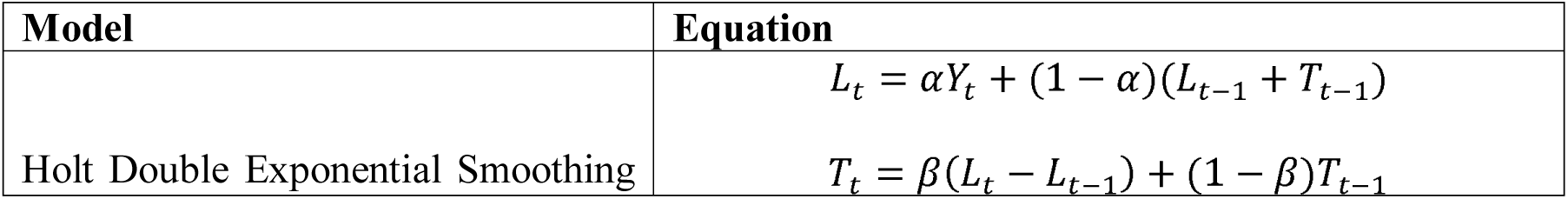
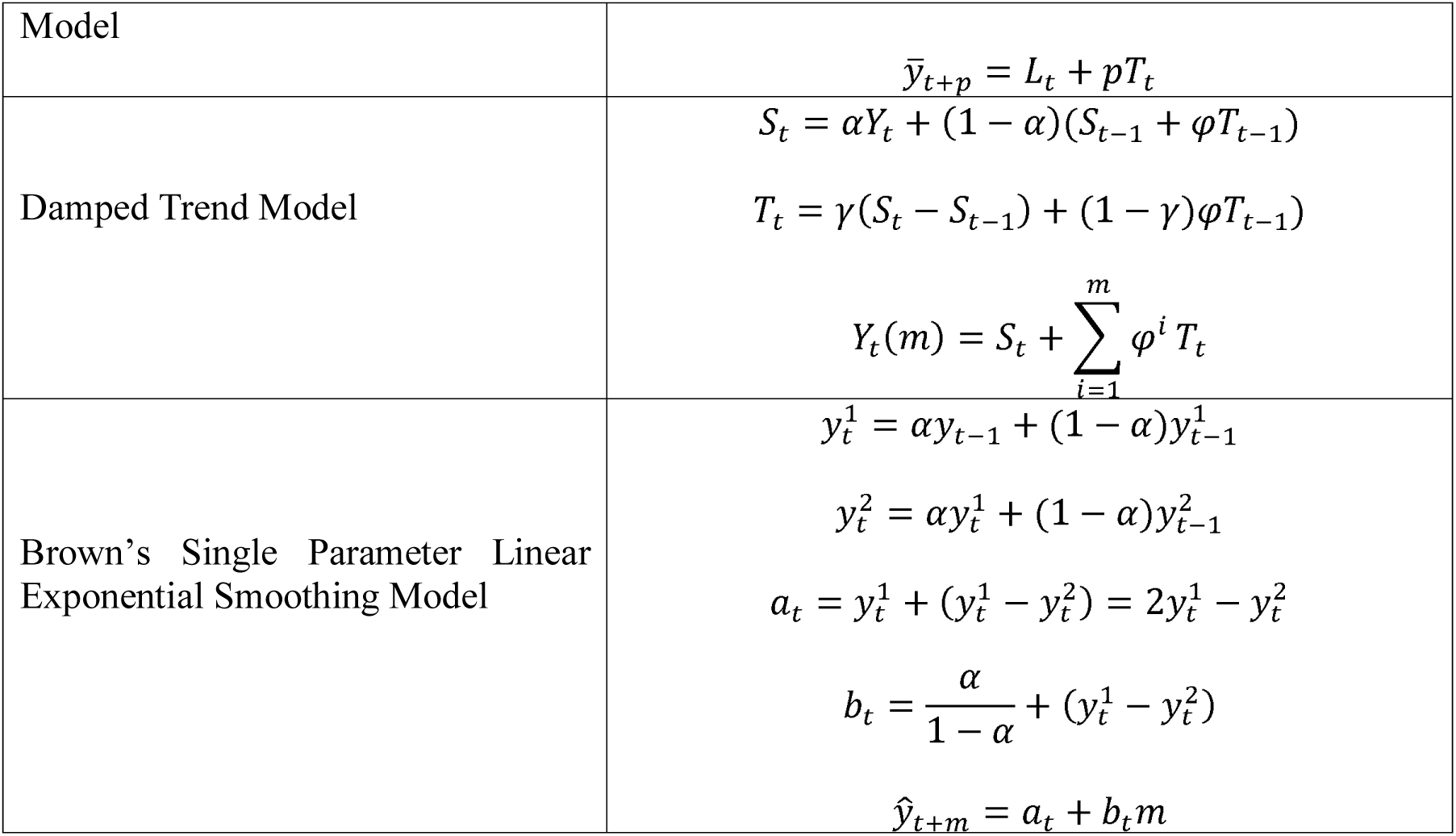
Exponential Smoothing Models

The models are examined in two subsections: first group models are Growth Curves and second group models are Time Series. The growth models descriptive equations are given in Table 1[3–8].

In Table 1, Y_t_ is the observed dependent variable as the number of total cases and the number of total death and ***t*** is the independent variable. In our models, ***t*** is the day. The ***A*** term is the is the asymptotic limit of the number of total cases and the number of total deaths as time goes to infinity, ***B*** is the proportion of the number of total cases to the number of total deaths. It is the proportion of the number of total cases to the number of total deaths, obtained after the estimated case / deaths with the initial value of time (days), to the highest number of total cases / deaths. The ***k*** term is the proportion of the maximum increase rate to the highest number of cases or death. The ***γ, c, d*** are the changing points that occurs when the change in the estimated increase rate goes from increase to decrease.

AR (p) is the p. degree of autoregressive series [9]. MA (q) refers to the moving average model of order *q*. In this series, *ε_t_~WN*(0,*σ*^2^) is the White noise series [10]. ARMA (*p, q*)) model is expressed by both AR (*p*) and MA (*q*) processes [11].

In Holt method, *L_t_* is the new smoothed value, *α* is the smoothing coefficient, (0<*α*<1), *Y* is the actual value at ***t***. period, *β* is the smoothing coefficient for trend estimation, (0<*β* < 1), *T_t_* is the trend predicted value, *p* is the number of forecasting periods and 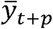 is the forecasting value after *p* period [12]. In Damped trend method, if 0<***φ***<1, the trend is damped, if *φ* = 1, the equations become identical to the Holt’s Linear Trend method. Tashman and Kruk (1996) defined that there may be value in allocating ***φ*** > 1, if applied in series with a strong tendency, with exponential trend [13]. The Brown’s Single Parameter Linear Exponential Smoothing Model is more suitable, if there is an increasing or decreasing trend in the time series. In this model, the initial equations 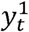 and 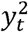 are obtained by single exponential smoothing and double exponential smoothing, respectively [14]. For the estimation of post m process, the equation is given in the below [15].

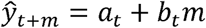

The exponential smoothing method is the method in which the estimates are constantly updated, taking into account the recent changes in the data [16]. In these methods, the weighted average of past period values is calculated and taken as the estimated value of future periods.

Estimation accuracy of the applied methods were evaluated with R^2^, Pseudo R^2^ and BIC. Bayesian information criterion (BIC) was developed by Gideon E. Schwarz (1978), who gave a Bayesian argument for adopting it [17].

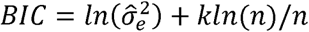

Where 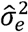 is the error variance

## Results

The data used in the study were the observations recorded between January 22 and April 21.

### Growth models prediction results for the number of total cases

The parameters estimates that estimated from 91 daily number of cases with nonlinear mathematical models between 22 January-15 April 2020 in China were presented in Table 4. Coefficient of determination (R^2^), Pseudo R^2^ and Mean Square Error (MSE) statistics were used to compare models. When Table 4 was examined, it was seen that the R^2^ and MSE values of Weibull, and Janoscheck models were equal. The MSE of Sloboda model was slightly smaller than two model but R^2^ was equal. The Sloboda model can be considered the most suitable model, through smaller MSE value, larger Pseudo R^2^ value. The Weibull and Janoscheck models can also be chosen as an alternative model.

**Table 4.** The parameter estimates and selection criteria results of growth models for the number of total cases

| Model | A | b | k | | MSE | $R^2$ |
| --- | --- | --- | --- | --- | --- | --- |
| Weibull | 81256.3 | 79123.8 | 0.000299 | $\gamma=2.706$ | 2609839.4 | 0.997 |
| Negatif exponential | 88954.9 |  | 0.042 |  | 72795574.9 | 0.903 |
| Von Bertalanffy | 82624.0 | 1.396 | 0.116 |  | 7835780.1 | 0.990 |
| Janoschek | 81256.3 | 0.974 | 0.000299 | $c=2.706$ | 2609839.4 | 0.997 |
| Lundqvist-Korf | 83881.5 | | 1814.408 | $d=-2.753$ | 10816583.1 | 0.986 |
| Sloboda | 81377.2 | 3.513 | 0.014 | $\gamma = 1.669$ | 2559449.5 | 0.997 |

The prediction curves of growth models are given in Figure 1.

**Figure 1.**
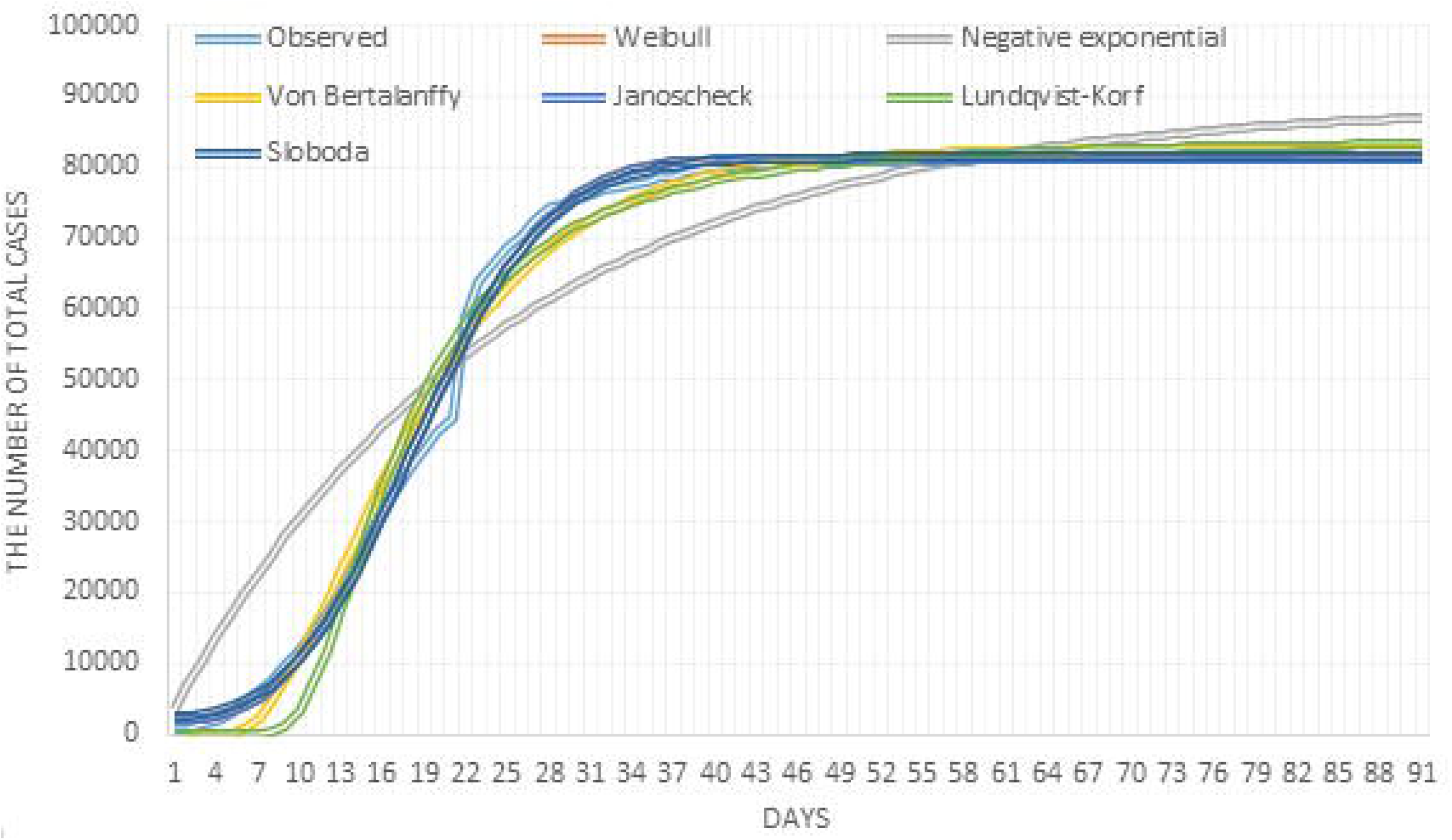
Estimation curves of growth models for the number of total cases

### The results of time series models for the number of total cases

Box-Jenkins methods and exponential smoothing methods were used among time series models for total number of cases. Autocorrelation (ACF) and partial autocorrelation (PACF) graphs of the series were examined. When the ACF and PACF graphs are examined in Figure 2, the first degree difference had been taken since the series were not stationary at the level. But the stationary assumption is not provided yet. The difference from the second degree was taken and the series became stationary. According to the ACF and PACF charts, the series quickly approached zero after the first delay in the ACF graph. In this case, since *p* = 0, d = 2 and *q* = 1, it was modeled by the integrated first degree moving averages method. In other words, the most suitable time series method was the ARIMA (0,2,1) model. In addition, exponential smoothing methods were used and the model performances were given in Table 5.

**Table 5.** Model compliance statistics for cases

| Model | R<sup>2</sup> | BIC | Ljung-Box |
| --- | --- | --- | --- |
| ARIMA(0,2,1) | 0.997 | 14.562 | 3.470 and p=0.999 |
| Holt | 0.997 | 14.600 | 3.925 and p=0.999 |
| Brown | 0.997 | 14.585 | 7.508 and p=0.976 |
| Damped | 0.997 | 14.639 | 3.480 and p=0.999 |

**Figure 2.**
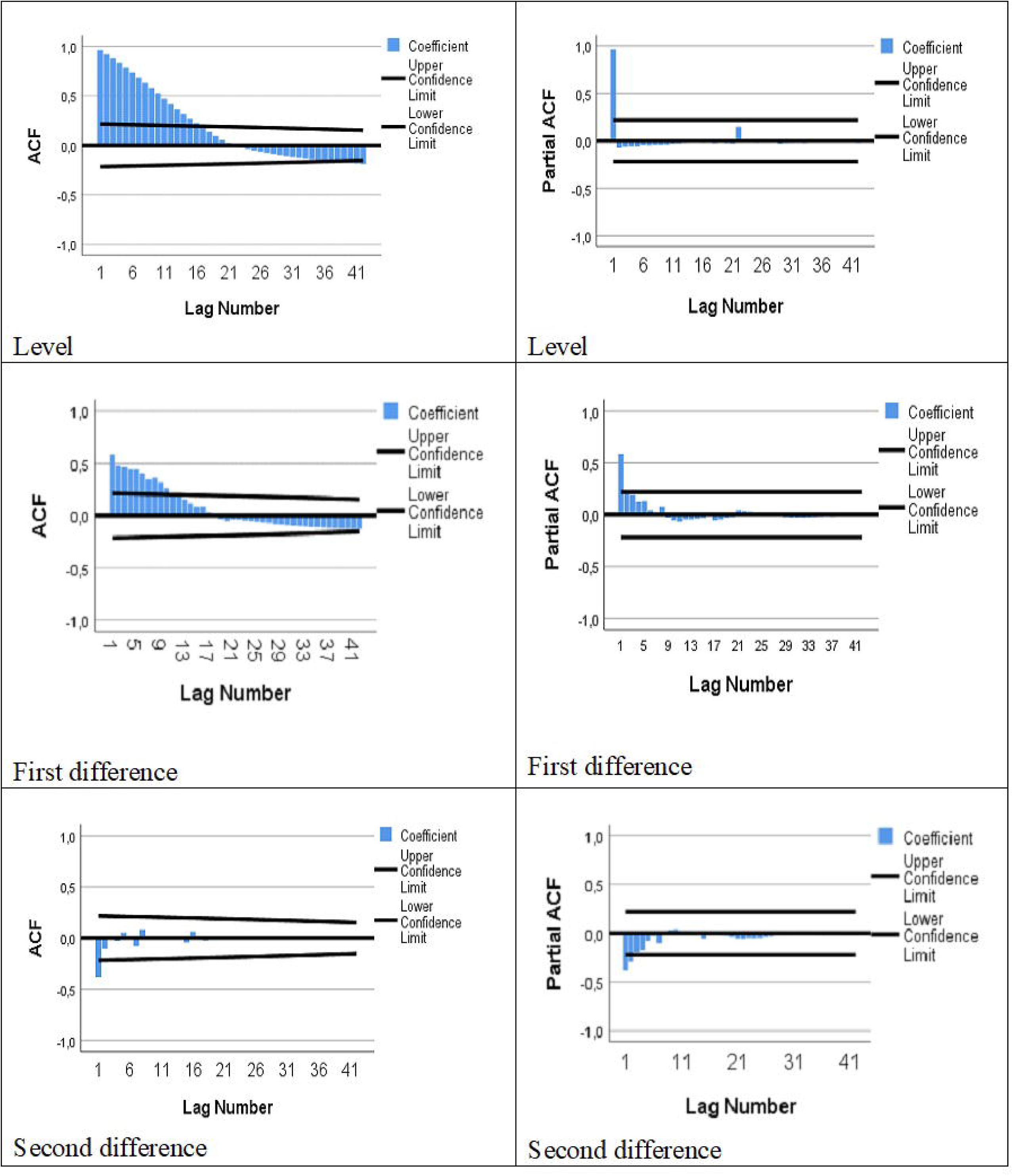
ACF and PACF graphs for cases

The performance of the model was given in Table 6 and it was seen that model’s estimations were successful like growth models.

**Table 6.** Agreement coefficients of ARIMA (0,2,1)

| Model Fit statistics |  |  |  | Ljung-Box Q |  |  |
| --- | --- | --- | --- | --- | --- | --- |
| Stationary R-squared | R-squared | RMSE | Normalized BIC | Statistics | DF | p |
| 0.266 | 0.997 | 1416.329 | 14.562 | 3.470 | 17 | 0.999 |

The parameter estimates of the ARIMA (0,2,1) model were given in Table 7.

**Table 7.**
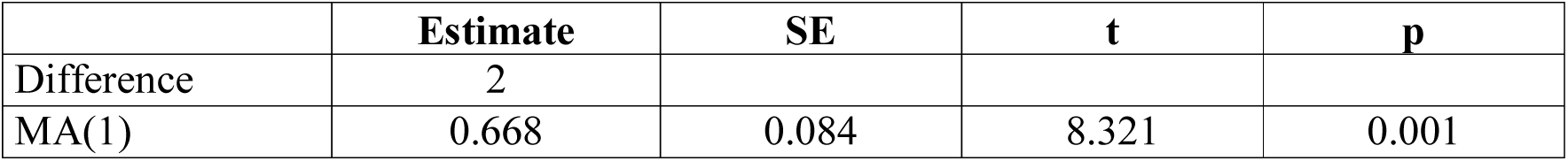
Model Parameters of ARIMA (0,2,1)

ARIMA (0,2,1) model was found appropriate among different time series models.

The ARIMA (0,2,1) model in this study can be written as in the below.

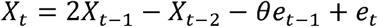

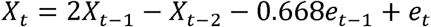

The forecasts from ARIMA (0,2,1) model for 15 day were given in Table 5.

**Table 5.** The predicted number of cases in China from April 22 to May 5

| Date (April 16- May 1 ) | Days | Case forecasting |
| --- | --- | --- |
| April 22 | 92 | 82798 |
| April 23 | 93 | 82837 |
| April 24 | 94 | 82877 |
| April 25 | 95 | 82916 |
| April 26 | 96 | 82955 |
| April 27 | 97 | 82995 |
| April 28 | 98 | 83035 |
| April 29 | 99 | 83074 |
| April 30 | 100 | 83114 |
| May 1 | 101 | 83153 |
| May 2 | 102 | 83193 |
| May 3 | 103 | 83232 |
| May 4 | 104 | 83272 |
| May 5 | 105 | 83311 |

As seen in Table 5, the number of total cases continues increasingly, albeit at low speed. The number of total cases is forecasting to be 83311 on the 105th day of the outbreak, that is, on 5 May 2020.

Observed and predicted values of the total cases were shown in Figure 3.

**Figure 3.**
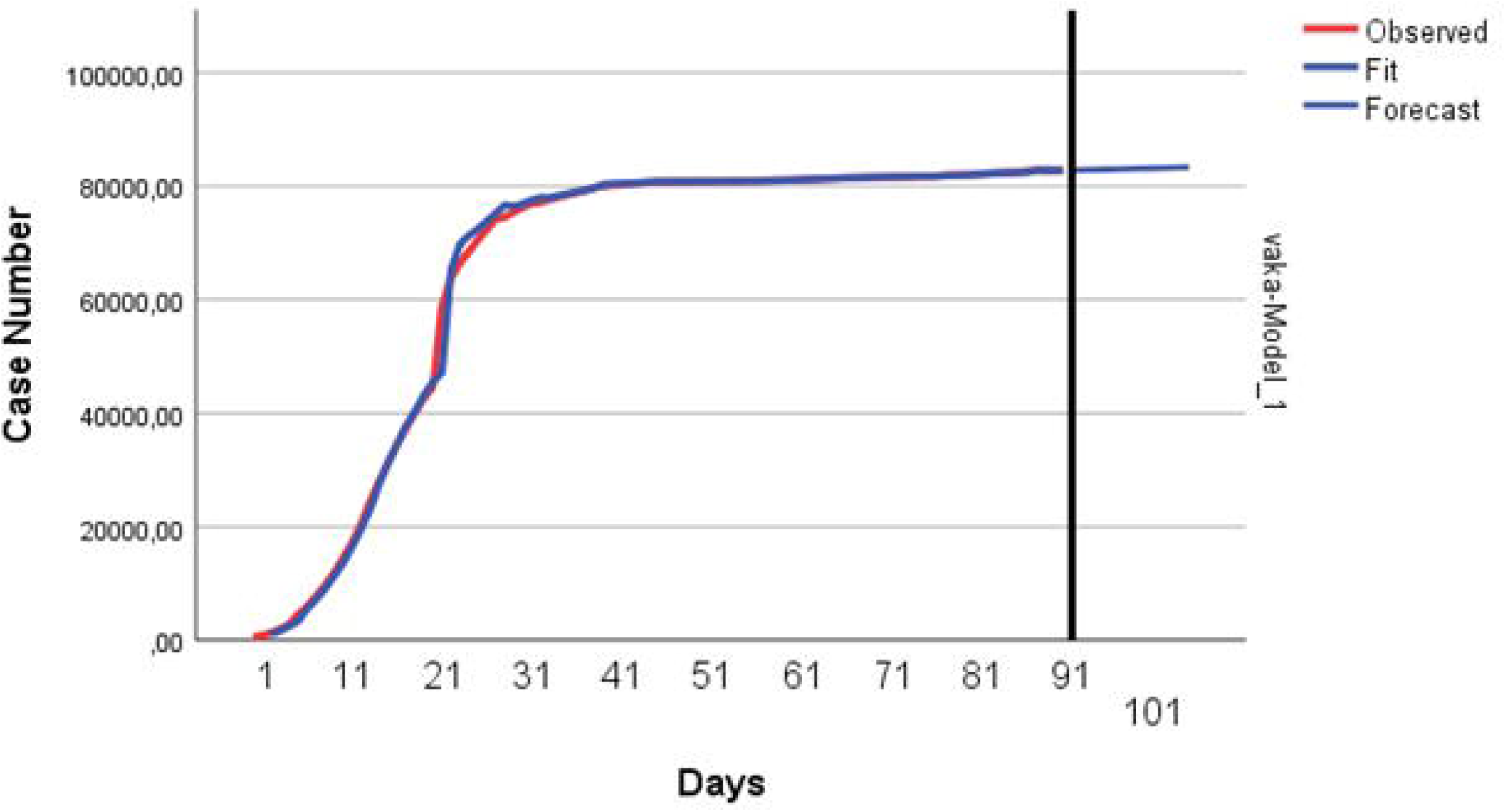
The curve of ARIMA(0,2,1) for the number of total cases

### The prediction and forecasting results of growth models for the number of total deaths

The parameters and the selection criteria of the growth models for total deaths were given in Table 8.

**Table 8.** The parameter estimates and selection criteria results of growth models for the number of total deaths

| Model | A | b | k |  | MSE | R<sup>2</sup> | Pseudo R<sup>2</sup> |
| --- | --- | --- | --- | --- | --- | --- | --- |
| Weibull | 3546.610 | 3631.067 | 0.002 | $\gamma=1.863$ | 88568.282 | 0.949 | Weibull |
| Negatif üstel | 5030.733 |  | 0.017 |  | 146041.563 | 0.913 | Negatif üstel |
| Von Bertalanffy | 3628.783 | 1.163 | 0.068 |  | 80308.717 | 0.953 | Von Bertalanffy |
| Janoschek | 3546.613 | 1.024 | 0.002 | $c=1.863$ | 88568.282 | 0.949 | Janoschek |
| Lundqvist-Korf | 4086.033 | | 194.275 | $d=-1.686$ | 75008.636 | 0.956 | Lundqvist-Korf |
| Sloboda | 3996.376 | 391039.638 | 8.721 | $\gamma=0.127$ | 76161.284 | 0.956 | Sloboda |

When Table 8 was examined regarding the number of total deaths in China, the most suitable models were Lundqvist-Korf and Sloboda growth models, respectively. The R^2^ values of these models were found highest as 0.956 and also MSE values were of these are lower than the others. Lundqvist-Korf model can be considered as the most suitable model, since it had smaller MSE. The prediction curves of growth models were given in Figure 4.

**Figure 4.**
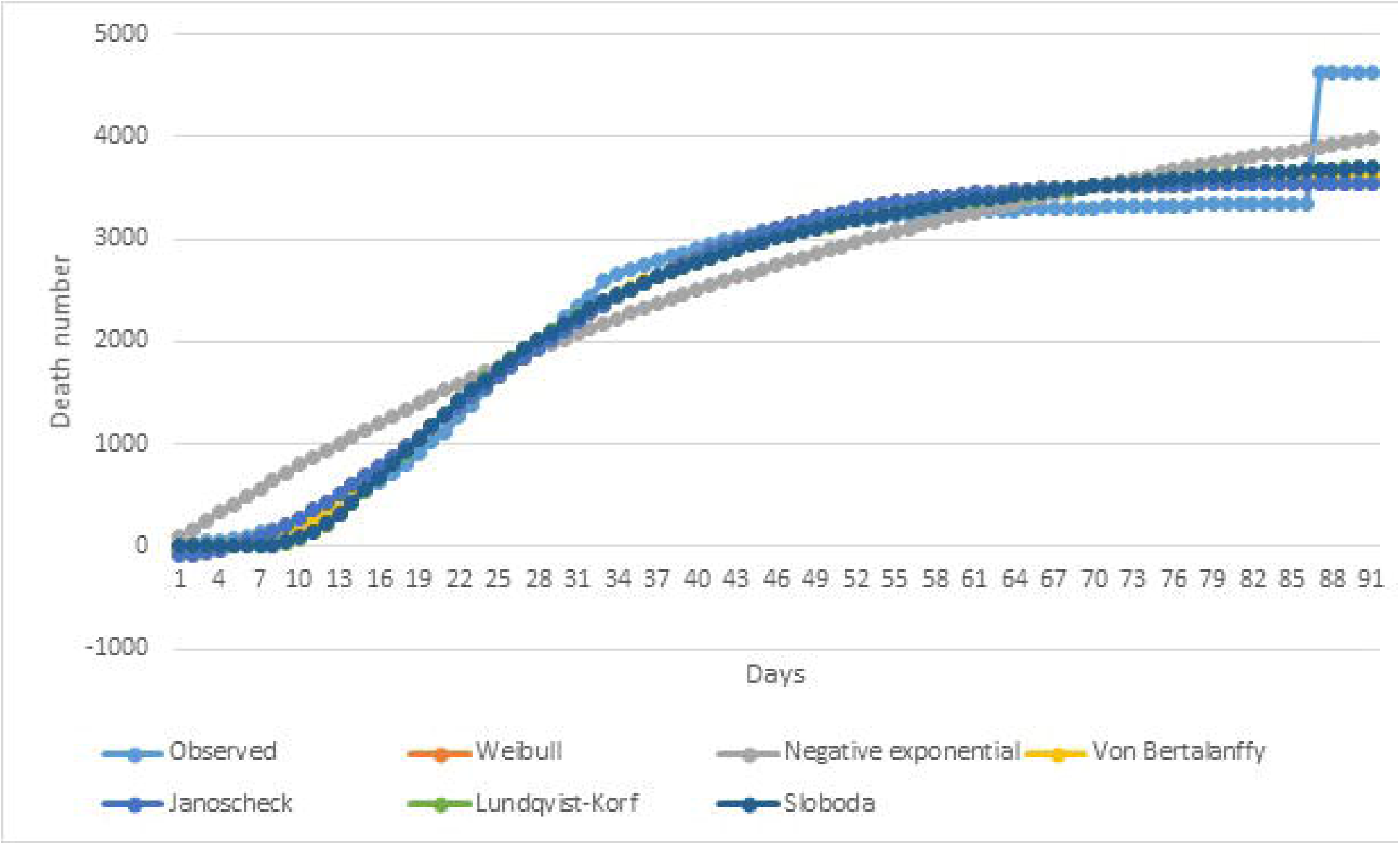
Curves of growth models for the number of total deaths

### The results of time series models for the number of total deaths

The most suitable time series model was found as Holt linear trend exponential smoothing model among time series models for the number of deaths. The compliance of the models were given in Table 9 and it was seen that the predictions were successful as growth models.

**Table 9.** Compliance statistics of the models

| Model | R<sup>2</sup> | BIC | Ljung-Box |
| --- | --- | --- | --- |
| ARIMA(1,2,0) | 0.982 | 10.372 | 12.609 and p=0.762 |
| Holt | 0.989 | 9.969 | 1.086 and p=0.999 |
| Brown | 0.987 | 10.036 | 11.590 and p=0.824 |
| Damped | 0.989 | 10.031 | 1.042 and p=0.999 |

The parameter estimation of the Holt linear trend exponential smoothing model was presented in Table 10. The observed and predicted values were given in Figure 5.

**Table 10.** The parameters of Holt linear trend exponential smoothing model

|  | Estimate | SE | t | Sig. |
| --- | --- | --- | --- | --- |
| Alpha (Level) | 1.000 | 0.107 | 9.388 | 0.001 |
| Gamma (Trend) | 0.001 | 0.015 | 0.054 | 0.957 |

SE: Standard error

The forecasts of the number of total deaths for exponential smoothing model by using Holt linear trend exponential smoothing model for 15 day were given in Table 11. The rate of increase in the number of deaths in China was decreasing and it was predicted that it will be between 3343-3355 in the period between April 16 and May 1, with a slight increase (Table 11).

**Table 11.**
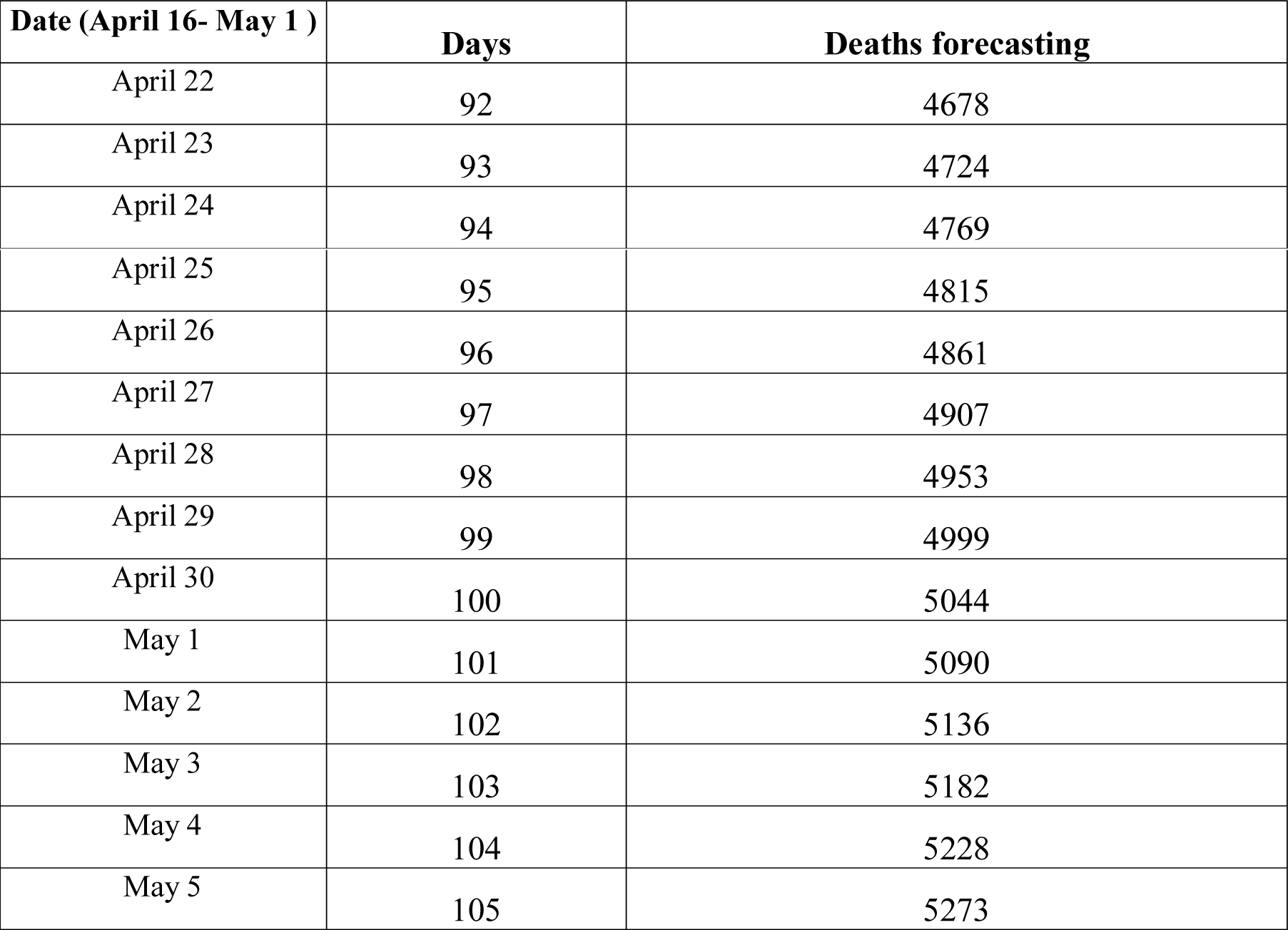
The forecasting results for Brown exponential smoothing model

The Holt linear trend exponential smoothing curve for the exponential smoothing model was given in Figure 5.

**Figure 5.**
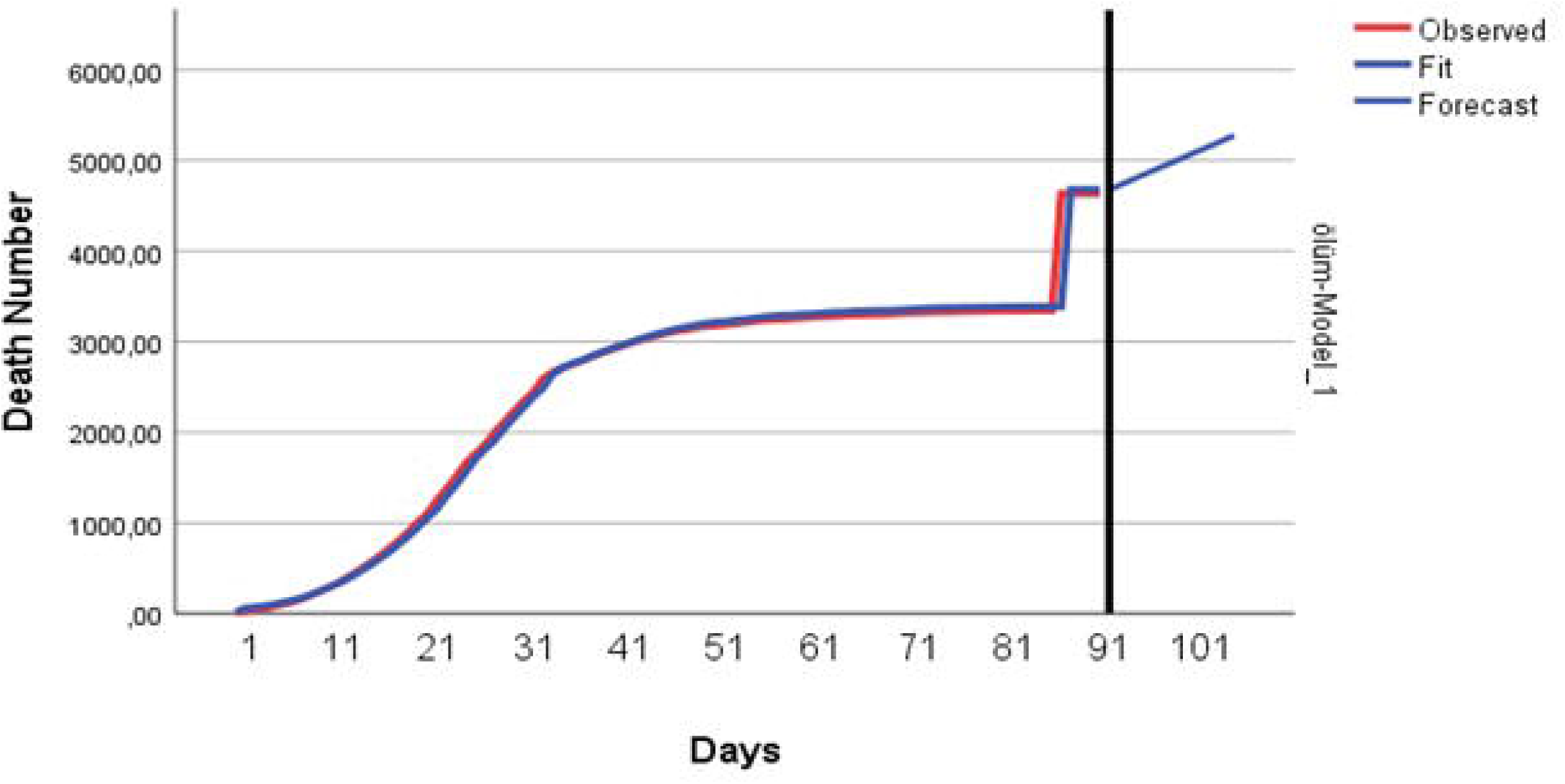
Curve of Holt linear trend exponential smoothing model for the number of total deaths

## Discussion

The best time series model was ARIMA (0,2,1) model for the number of cases. In a study, ARIMA model was used on the daily prevalence data of COVID-2019 from January 20, 2020 to February 10, 2020 and ARIMA (1,2,0) and ARIMA (1,0,4) models were obtained [18]. Logistic, Bertalanffy and Gompertz models were used to estimate the number of cases and deaths on COVID-19 disease in different regions in China before by Jia and et al. According to their study, the Logistics model was reported to be better than the others by the R^2^ criterion conducted extensive research in quasi-experimental analysis method in various provinces in China and investigated the relationship between population and number of outbreak cases [19,20]. In their study, they found that the correlation coefficients of the relationship between the population and the number of cases differed among regions. They have observed that the number of cases was high in regions with high population and there was a high correlation between them. They stated that factors such as immigrants, tourism and mobility plays an important role in this. Also the authors determined the number of cases with the epidemic growth model Fan et al. [19].

Roosa et al. (2020) analyzed the number of cases in some regions in China with generalized logistic growth model (GLM), Richards Model and Sub-Epidemic Model in a short-term (10 days). They found that the number of cases will increase. They estimated that the case increase (GLM) in the Guangdong and Zhejiang regions would be lower by using the Richards models and the rate would be higher by using the sub-epidemic model [21].

In a study on the risk of infection of COVID-19 detected in a passenger ship in China in February 2020, it was observed that the risk of infection in those with close contact was higher than those with no close contact. The estimated of number of cases was obtained by back calculation method [22]. Al-qaness et al. (2020) used Adaptive Neuro-Fuzzy Inference System (ANFIS), Flower Pollination Algorithm (FPA), Salp Swarm Algorithm (SSA) and FPASSA-ANFIS methods to estimate the number of cases with COVID-19 disease in China and the USA. They calculated model performance with Root Mean Square Error (RMSE), Mean Absolute Error (MAE), Mean Absolute Percentage Error (MAPE), Root Mean Squared Relative Error (RMSRE) and R^2^. They found that the best method for modeling and estimating the number of total cases was the FPASSA-ANFIS method [23].

Kuniya (2020) estimated the outbreak peak of coronavirus disease in Japan with the SEIR compartmental model [24]. In another study, the number of reproduction of the Wuhan novel coronavirus 2019-nCoV was estimated with the susceptible-exposed-infected-removed (SEIR) compartment model [25]. There are studies on coronavirus disease by different researchers using various statistical methods. Yuan et al. Used the median (interquartile range, IQR) and Mann Whitney U test or Wilcoxon test, Twu et al and Prem et al. Used SEIR model and Neher et al used SIR model [26–29]. In our study, we compared the time series analysis using Weibull, Negative Exponential, Von Bertalanffy, Janoscheck, Lundqvist-Korf and Sloboda models, which are different from the methods used in previous studies. According to the literature, there is no publication about nonlinear mathematical models used in our study on coronavirus outbreak before.

## 5. Conclusions

The parameter estimates of the Weibull, Janoschek, Sloboda and Lundqvist-Korf were close to each other in the analysis of nonlinear growth models regarding the number of cases and deaths in China. Their R^2^ and MKE statistics were similar. The parameter estimation and compliance statistics of the Negative Exponential and von Bertalanffy models, which are among the nonlinear models, differed in both the number of cases and the number of deaths. When nonlinear models are examined, R^2^ value, which is used as a criterion in comparing models, was obtained from the lowest Negative Exponential model both in the number of total cases and the number of total deaths. The lowest MSE was obtained from Sloboda model for the number of total cases and from Lundqvist-Korf model for the number of total deaths.

In terms of time series analysis, the number of total cases and the number of deaths are modeled differently. The number of total cases were modelled with ARIMA(0,2,1) that is a moving averages method, while the number of total deaths were modeled by the Holt linear trend model, which is a exponential smoothing method. According to the estimation results, we estimate that the number of total cases and deaths will be increase and this will be a big danger. For this result, all necessary precautions must be taken against the danger.

## Data Availability

https://www.who.int/news-room/detail/08-04-2020-who-timeline---covid-19

## References

1. WHO Timeline - COVID-19. Available online: https://www.who.int/news-room/detail/08-04-2020-who-timeline---covid-19 (accessed on 20 April 2020).

2. Panik, M. J. Growth Curve Modeling. Theory and Applications. 1^st^ ed.; John Wiley and Sons, Inc., Hoboken, New Jersey, Canada, 2014; pp.437.

3. von Bertalanffy, L. Quantitative Laws in Metabolism and Growth. Quarterly Review of Biology 1957, 32, 217–231.

4. Korf, V. A. Mathematical Definition of Stand Volume Growth Law. Lesnicka Prace 1939, 18, 337–339.

5. Lundqvist, B. On the Height Growth in Cultivated Stands of Pine and Spruce in Northern Sweden. Meddelanden Fran Statens Skogsforsknings-institut 1957, 47, 1–64.

6. Sloboda, B. Investigation of Growth Processes Using First-Order Differential Equations. Mitteilungen der Baden-Württembergischen Foustlichen Versuchs und Forschungsanstalt. Heft 1971a, 32.

7. Sloboda, B. Zur Darstellung von Washstumprozessen mit Hilfe von Differentialgleichungen evster Ordung. Mitteilungen der Baden-Württembergischen Foustlichen Versuchs und Forschungsanstalt. 1^st^ ed.; Baden-Württemberg: Baden-Württembergische Forstliche Versuchsund Forschungsanstalt. 1971b; pp.1.

8. Weibull, W. A. Statistical Distribution Function of Wide Applicability. Journal of Applied Mechanics 1951,18, 291–297.

9. Wei, W. W. S. Time Series Analysis. 2^nd^ ed.; Addison Wesley Publishing Company, New York, 2006; pp. 156.

10. Montgomery, D. C.; Johnson, L. A.; Gardiner, J. S. Forecasting and Time Series Analysis, 1^st^ ed.;McGraw-Hill, Inc., USA, 1990; pp.249.

11. Cryer, J. D. Time Series Analysis, 1^st^ ed.; PWS Publishers, USA, 1986; pp.89.

12. Holt, C. C. Forecasting seasonals and trends by exponentially weighted moving averages. International Journal of Forecasting 2000, 20, 5–5.

13. Tashman, L.; Kruk, J. The use of protocols to select exponential smoothing procedures: a reconsideration of forecasting competitions. International Journal of Forecasting 1996, 12,235–18.

14. Armutlu, I. H. İşletmelerde Uygulamalı İstatistik Sayısal Yöntemler-1. 2^nd^ ed.;Alfa Yayınları, 2. Baskı, İstanbul, Turkey, 2008; pp.1

15. Orhunbilge, N. Zaman Serileri Analizi Tahmin ve Fiyat Endeksleri, 1^st^ ed.; Avcıol BasımYayın, İstanbul, Turkey, 1999; pp.1

16. Kadılar, C. SPSS Uygulamalı Zaman Serileri Analizine Giriş, 1^st^ ed.; Bizim Büro Basımevi, Ankara, Turkey, 2009; pp.1

17. Schwarz, G. E. Estimating the dimension of a Model. Annals of Statistics 1978, 6, 461–464.

18. Benvenuto, D.; Giovanetti, M.; Vassallo, L.; Angeletti, S.; Ciccozzi, M. Application of the ARIMA model on the COVID-2019 epidemic dataset. Data in brief 2020, 29, 105340.

19. Fan, C.; Liu, L.; Guo, W.; Yang, A.; Ye, C.; Jilili, M.; Ren, M.; Xu, P.; Long, H.; Wang, Y. Prediction of Epidemic Spread of the 2019 Novel Coronavirus Driven by Spring Festival Transportation in China: A Population-Based Study. International Journal of Environmential Research and Public Health 2020, 17, 1679.

20. Jia, L.; Li, K.; Jiang, Y.; Guo, X.; Zhao, T. Prediction and analysis of Coronavirus Disease 2019. Quantitative Biology 2020, arXiv preprint arXiv:2003.05447, 2020.

21. Roosa, K.; Lee, Y.; Luo, R.; Kirpich, A.; Rothenberg, R.; Hyman, J. M.; Yan, P.; Chowell, G. Short-term Forecasts of the COVID-19 Epidemic in Guangdong and Zhejiang, China: February 13–23, 2020. Journal of Clinical Medicine 2020,9, 596.

22. Nishiura, H. Backcalculating the Incidence of Infection with COVID-19 on the Diamond Princess. Journal of Clinical Medicine 2020, 9, 657.

23. Al-qaness, M. A. A.; Ewees, A. A.; Fan, H.; Abd El Aziz, M. Optimization Method for Forecasting Confirmed Cases of COVID-19 in China. Journal of Clinical Medicine 2020, 9, 674.

24. Kuniya, T. Prediction of the Epidemic Peak of Coronavirus Disease in Japan, 2020. Journal of Clinical Medicine, 2020, 9, 789.

25. Zhou, T.; Liu, Q.; Yang, Z.; Liao, J.; Yang, K.; Bai, W.; Lu, X., Zhang, W. Preliminary prediction of the basic reproduction number of the Wuhan novel coronavirus 2019-nCoV. J Evid. Based. Med. 2020, 13, 3–7.

26. Yuan, M.; Yin, W.; Tao, Z.; Tan, W., Hu, Y. Association of radiologic findings with mortality of patients infected with 2019 novel coronavirus in Wuhan, China. Plos One 2020,15: e0230548. https://doi.org/10.1371/journal.pone.0230548.

27. TWu, J.; Leung, K.; Leung, G. M. Nowcasting and forecasting the potential domestic and international spread of the 2019-nCoV outbreak originating in Wuhan, China: a modelling study. Lancet 2020, 395, 689.

28. Prem, K.; Liu, Y.; Russell, T. W.; Kucharski, A. J.; Eggo, R. M.; Davies, N. The effect of control strategies to reduce social mixing on outcomes of the COVID-19 epidemic in Wuhan, China: a modelling study. Lancet Public Health 2020, https://doi.org/10.1016/S2468-2667(20)30073-6.

29. Neher, R. A.; Dyrdak, R.; Druelle V.; Hodcroft, E. B.; Albert, J. Potential impact of seasonal forcing on a SARS-CoV-2 pandemic. Swiss Medical Weekly 2020,150, w20224.

